# Chest Computed Tomography Findings in Asymptomatic Patients with COVID-19

**DOI:** 10.1101/2020.05.09.20096370

**Authors:** Min Cheol Chang, Wonho Lee, Jian Hur, Donghwi Park

## Abstract

**Background:** Little is known about the damage to the respiratory system in asymptomatic patients with coronavirus disease (COVID-19).

**Objective:** Herein, we evaluated the findings of chest computed tomography (CT) and radiography in patients with COVID-19 who were asymptomatic.

**Methods:** We retrospectively investigated patients with a confirmed diagnosis of COVID-19 but who did not show any symptoms. Among the 139 patients with COVID-19 who were hospitalized, 10 (7.2%) were asymptomatic. Their chest CT and radiographic findings were analyzed.

**Results:** In the results, all patients (100%) had ground glass opacity (GGO) on chest CT. Further, the GGO lesions were predominantly distributed peripherally and posteriorly in all patients. In 9 (90%) patients, the GGO lesions were combined with reticular opacity. Air-bronchogram due to bronchiolectasis surrounded by GGO was observed in 8 patients (80%). Additionally, the lung lesions were dominant on the right side in all patients.

**Conclusions:** In conclusion, considering our results that the lung is affected in asymptomatic patients, it will be necessary to extend the indications of COVID-19 testing for effective management of COVID-19 during the pandemic.

## Introduction

In December 2019, an outbreak of the coronavirus disease (COVID-19) occurred in Wuhan, China, and the disease subsequently spread to approximately 200 countries within 3 months. [1, 2] This disease causes massive alveolar damage and progressive respiratory failure, with 2-6% of COVID-19 cases leading to mortality. [3] The fatality rate is higher in older individuals and individuals having underlying diseases. Because COVID-19 is highly infectious, individuals who come in close contact with COVID-19 patients and those who have a confirmed diagnosis of COVID-19 are placed in isolation to prevent secondary and tertiary infections. Typical symptoms of COVID-19 include fever, cough, sputum, chest pain, and muscle soreness. However, approximately 1-5% of patients with COVID-19 are reported to be asymptomatic. [4-6] Asymptomatic COVID-19 patients are likely not to be tested and will remain unaware of the fact that they have infection. Therefore, asymptomatic patients with COVID-19 can be a source for the spread. Moreover, if asymptomatic patients are left untreated, then damage to the respiratory system may occur. However, little is known about the damage to the respiratory system in asymptomatic COVID-19 patients. Therefore, in the current study, we evaluated the findings of chest computed tomography (CT) and radiography in patients with COVID-19 who were asymptomatic.

## Methods

### Patients

This is a retrospective study. Of 139 confirmed COVID-19 patients admitted to our university hospital (a 930-bed, tertiary referral hospital) between February 18, 2020, and April 11, 2020, we selected patients with confirmed COVID-19 who did not show any symptoms of COVID-19 at the initial diagnosis and during the treatment period. All included patients had their diagnosis confirmed through RT-PCR (Allplex™ 2019-nCoV Assay, Seegene®, South Korea) with a pharyngeal swab at our hospital.

In all patients who were investigated in this study, chest CT and radiography were performed as a standard process during admission to our hospital. This study was approved by the Institutional Review Board of a University Hospital.

### Chest CT Protocol

All images were obtained using a CT system (Siemens Healthineers, Germany) with patients in the supine position. The main scanning parameters were as follows: tube voltage = 120 kVp, automatic tube current modulation (30-70 mAs), pitch = 0.99-1.22 mm, matrix = 512 × 512, slice thickness = 10 mm, and field of view = 350 mm × 350 mm. All images were then reconstructed with a slice thickness of 0.625-1.250 mm with the same increment.[7]

### Qualitative image analysis

One radiologist (WL), with 25 years of experience, checked and confirmed the radiologic findings. We reviewed the initial chest CT images and radiographies taken on admission for each patient. We checked the CT findings for the same parameters as those mentioned in the previously reported cases: number of affected lobes, presence of GGO, consolidation, halo or reversed halo signs, crazy paving pattern, air-bronchogram manifested as bronchiolectasis surrounded by GGO, reticular opacity suggesting interstitial thickening or fibrosis, subpleural line, and other combined pathologies including lymphadenopathy and pleural effusion.[7]

## Results

### Patients ’ characteristics

Among the 139 patients with COVID-19 who were hospitalized, 10 (7.2%) were asymptomatic. Their mean age was 65 ± 12.8 years (age range: 52-95 years) and the sex ratio was 6:4 (male:female)(Table 1). Five patients (50%) underwent tests for COVID-19 because they had a contact history or possibility of contact with COVID-19 patients, and 2 patients (20%) had COVID-19 tests as a routine laboratory test on admission for treating other underlying disorders. The other 3 patients (30%) received COVID-19 tests as a routine laboratory test before a surgical operation. Our patients were treated with either hydroxychloroquine sulfate (Oxiklorine®; 400 mg qd per day) alone or a combination of hydroxychloroquine sulfate (Oxiklorine®; 400 mg) and lopinavir/ritonavir (400 mg/100 mg bid per day; Kaletra®). All 10 patients were admitted for at least 2 weeks, and did not show any symptoms of COVID-19 until they were discharged to home after cured.

### Findings of Computed Tomography

The findings of chest CT and radiography in each patient are summarized in Table 1. Representative CT and radiographic images of each patient are presented in Figure 1. In the 10 asymptomatic patients, the mean number of affected lobes was 2.4 ± 1.3. Three (30%) patients had 1 affected lobe, 2 (20%) patients had 2 affected lobes, 4 (40%) patients had 3 affected lobes, while 1 (10%) patient had 5 affected lobes.

**Figure 1.**
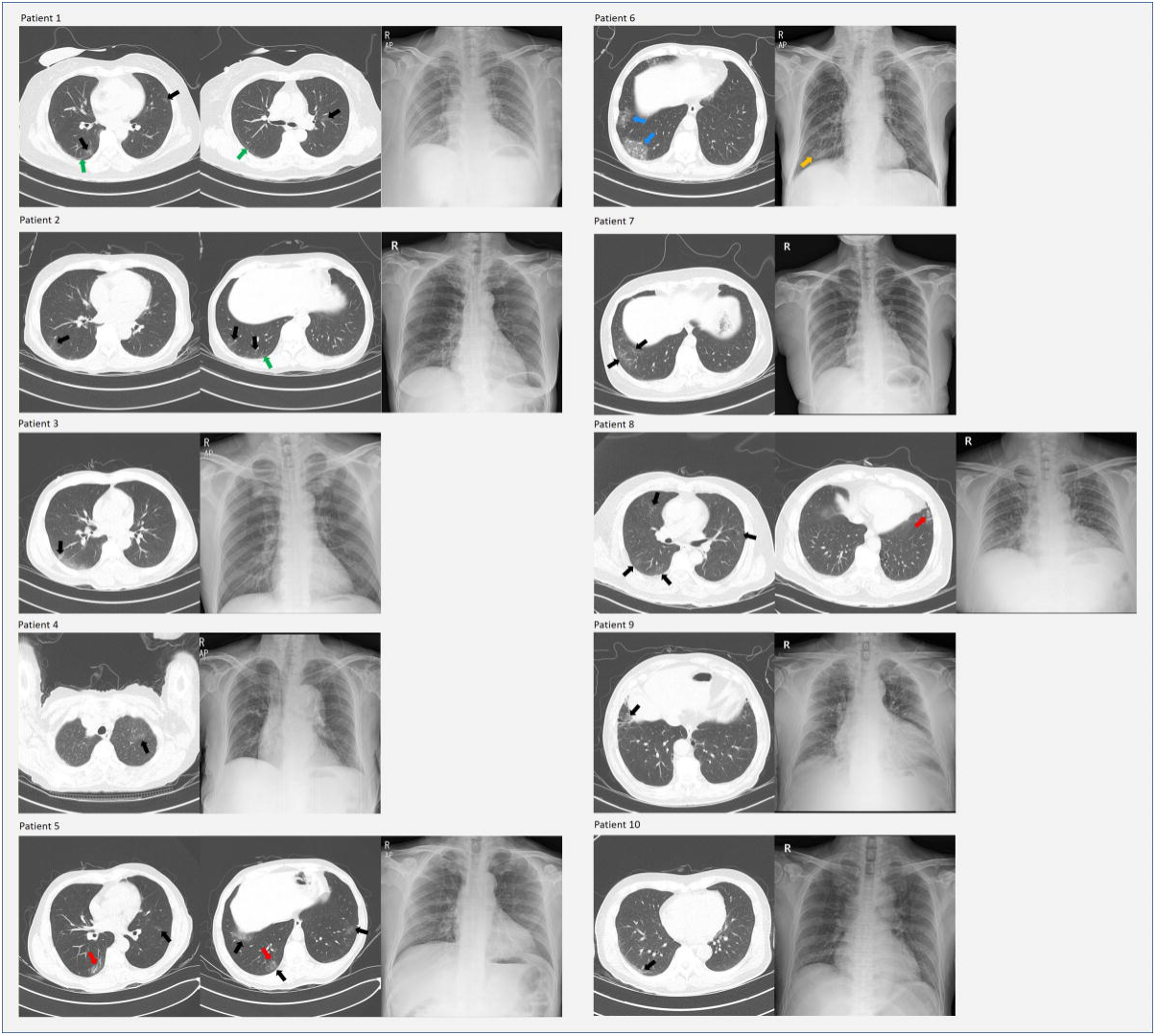
Chest computed tomographic and radiographic images of each patient. Black arrow: ground glass opacity (GGO) and/or reticular opacity. Red arrow: Air-bronchogram (bronchiolectasis with surrounding GGO and/or reticular opacity). Blue arrow: Crazy paving appearance. Green arrow: Subpleural linear opacity. Orange arrow: GGO in chest radiograph

Regarding chest CT findings, in all 10 asymptomatic patients, findings of consolidation and halo or reversed halo signs were not observed. All patients (100%) showed GGO on chest CT. Further, the GGO lesions were predominantly distributed peripherally and posteriorly in all patients. The GGO lesions of 9 (90%) patients were combined with reticular opacity of intralobular septal thickening, and air-bronchogram due to bronchiolectasis surrounded by GGO was observed in 8 patients (80%). A definite crazy paving pattern appeared in only 1 patient (10%). Three patients (30%) showed subpleural lines. Additionally, the lung lesions were dominant on the right side in all patients (Table 2).

### Findings of Chest Radiographs

On chest radiographs, 1 patient (10%) had subtle GGO. The other 9 patients’ radiographs revealed no specific abnormal findings.

## Discussion

Among the 139 patients with COVID-19 who were hospitalized, 10 (7.2%) were asymptomatic. In previous reports, 4%–25% of people testing positive for COVID-19 were reported to be asymptomatic.[8-10] The widely heterogeneous percentage of asymptomatic patients in previous studies may be due to different support systems for the cost of COVID-19 testing and different policies on screening in each country.

In the results of this study, unlike chest radiography, where only 10% were positive, all asymptomatic patients showed abnormal findings on their CT scans. Considering these false-negative results of chest radiography, CT scans must be recommended to confirm lung involvement in asymptomatic patients with COVID-19. In previous studies, 83.9–88.7% of symptomatic COVID-19 patients showed abnormal CT findings.[10-12] In this study, however, all patients showed abnormal CT findings despite no symptoms. Our results may be due to the fact that we did not check the chest CT immediately after diagnosis but a few days later after confirmation of COVID-19 (7.8 ± 5.6 days). We hypothesize that the SARS-CoV-2 was not allowed enough time to damage the patients’ lung in previous studies.

In this study, the CT scans showed increasing involvement of the peripheral and posterior distributions in asymptomatic patients with COVID-19, which is consistent with the CT findings of a previous study in symptomatic patients with COVID-19.[10, 11] Zhou et al. [10] reported that lesions had a characteristic multifocal distribution in the middle and lower lung regions and in the posterior lung area as noted in the CT scan of symptomatic patients with COVID-19. Guan et al. [11] reported that more than 70% of CT scans in symptomatic patients with COVID-19 showed lesions in the lower lobe. In all our patients, the right lung was predominantly involved. This finding was also consistent with that reported in a previous study. [10] Zhou et al. [10] reported that, of the 10 patients with a single lesion, 9 (90.0%) had lesions that were initially located in the right lung; and one (10.0%) had a lesion in the left lung. These results may be due to the innate anatomic features of the right inferior lobar bronchus. The bronchus of the right lower lobe of the lung is steeper and straighter than the left bronchial branches, and the angle between the right lower lobe and the long axis of the trachea is smaller; hence, in the early phase of COVID-19, the virus is more likely to invade the branches of the right inferior lobar bronchus and cause infection. [10]

Regarding the change in CT findings according to the progression of COVID-19, Zhou et al. [10] showed the differences between abnormal CT findings in the early phase (≤7 days after symptom onset) and those in the advanced phase (8–14 days after symptom onset) of COVID-19 in 62 patients with COVID-19. In their study, they observed many GGO findings in the early phase (47.5% vs. 27.3%) and then an increase in GGO with reticular opacity (50.0% vs. 86.4%) and air-bronchogram (62.5% vs. 90.9%) in the advanced phase (4). Considering these results, in our study, the incidence rate of GGOs with reticular opacity and air-bronchograms was 90% and 80%, respectively; accordingly, CT patterns were somewhat similar to those in the advanced stage despite being asymptomatic.

Our study has shown that asymptomatic COVID-19 patients also suffer significant lung damage. Although the prognosis of untreated patients with COVID-19 has not been reported, our results show that active screening may be necessary to detect asymptomatic patients with COVID-19 and manage them appropriately. Our study is the first to analyze the CT findings of asymptomatic patients with COVID-19. However, the present study has two limitations. First, changes in CT over the course of COVID-19 pneumonia have not been fully tracked and described for all patients. Second, we recruited a relatively small number of subjects. In the future, further studies to compensate for our limitations are required.

## Funds

The present study was supported by a National Research Foundation of Korea grant funded by the Korean government (grant no. NRF-2019M3E5D1A02068106)

## Authorship contribution statement

Min Cheol Chang: Data curation, Writing - original draft. Jian Hur: Data curation, Conceptualization, Methodology. Donghwi Park: Conceptualization, Methodology, Writing - review & editing, Supervision. Wonho Lee: Data curation,

## Acknowledgement

None

## Statement of Ethics

Ethical approval was obtained from the IRB of the Yeungnam University Hospital.

## Data Availability

Data will be provided if there is reasonable request.

